# Risk of Incident Atrial Fibrillation in Women with a History of Hypertensive Disorders of Pregnancy: A Population-Based Retrospective Cohort Study

**DOI:** 10.1101/2024.10.11.24315357

**Authors:** Amy Johnston, William Petrcich, Graeme N. Smith, Deshayne B. Fell, Peter Tanuseputro, Thais Coutinho, Jodi D. Edwards

## Abstract

**Background:** Hypertensive disorders of pregnancy (HDP) are a major cause of maternal morbidity and mortality and are associated with acute cardiac events in the peripartum period, as well as cardiovascular disease (CVD) later in life. Despite the robust association between hypertension and atrial fibrillation (AFib), comparatively little is known about HDP and individual HDP subtypes as sex-specific risk factors for AFib.

**Methods:** A population-based retrospective cohort study of 771,521 nulliparous women discharged for obstetrical delivery of their first live or stillborn singleton infant between 2002-2017 in Ontario, Canada. Data were obtained from record-level, coded, and linked population-based administrative databases housed at ICES. Using competing risks Cox proportional hazards regression, we estimated crude and multivariable-adjusted cause-specific hazard ratios (csHRs) and 95% confidence intervals (CIs) for associations between history of any HDP–and its six subtypes–and AFib before death, as well as all-cause mortality without a prior AFib diagnosis.

**Results:** Approximately 8% of subjects were diagnosed with HDP during the 16-year exposure accrual period. The total person-time of follow-up was 7,380,304 person-years, during which there were 2,483 (0.3%) incident AFib diagnoses and 2,951 (0.4%) deaths. History of any HDP was associated with an increased csHazard of both incident AFib and death without a prior AFib diagnosis [adjusted csHRs (95% CIs): 1.45 (1.28-1.64) and 1.31 (1.16-1.47), respectively]. These associations were observed in relatively young women (median time-to-event: 7 years postpartum). Associations suggestive of a ‘dose-response’ relationship were also observed, whereby both HDP severity, and presence of pre-pregnancy chronic hypertension, were associated with higher rates of both outcomes.

**Conclusions:** People exposed to HDP in their first delivery have a significantly increased csHazard of incident AFib compared to their unexposed counterparts, with higher rates observed in subjects exposed to more severe *de novo* HDP diagnoses as well as chronic hypertension in pregnancy. Given the substantial morbidity and mortality burden of AFib in women, these findings underscore the critical importance of considering history of HDP in risk calculation/stratification for both arrhythmic and non-arrhythmic CVDs; improving population-based surveillance of traditional and female-specific CVD risk factors; and developing targeted prevention strategies aimed at reducing the occurrence and burden of HDP.

**Clinical Perspective What is new?:**

- In this population-based retrospective cohort study of 771,521 nulliparous women, a history of hypertensive disorders of pregnancy (HDP) significantly increased the cause-specific hazard of incident atrial fibrillation (AFib) compared to women without HDP, even after adjustment for confounders, and this association was observed in relatively young women (median follow-up: 7 years postpartum).
- Associations suggestive of a ‘dose-response’ relationships were observed, whereby subjects with more severe *de novo* HDP diagnoses, as well as those with pre-pregnancy chronic hypertension, had higher cause-specific rates of AFib, with the highest rate observed in subjects exposed to chronic hypertension in pregnancy.

**What are the clinical implications?:**

- These findings suggest that women with a history of any HDP–especially those with pre-pregnancy chronic hypertension–may benefit from closer monitoring for the early detection of AFib.
- Enhanced population-based surveillance of, and targeted strategies to prevent, HDP as a female-specific cardiovascular risk factor are needed to mitigate intermediate- and long-term cardiovascular disease risk associated with these adverse pregnancy conditions.

## Introduction

Hypertensive disorders of pregnancy (HDP), which affect between 5 to 15% of all pregnancies,^1, 2^ are a heterogeneous group of obstetrical complications with differing epidemiology, pathophysiology, and symptomatology.^3^ They are a major cause of maternal and fetal/neonatal morbidity and mortality,^4^ with several studies reporting associations between a history of HDP and acute cardiac events in the peripartum period, as well as cardiovascular disease (CVD), later in life.^5, 6^

The pathophysiological mechanisms linking HDP and CVD remain unclear.^7^ However, observed associations may be driven by an interplay of several pregnancy-related factors that act through both maternal (e.g., advanced maternal age, history of obstetrical complications) and placental pathways, combined with additional accumulated exposure to non-pregnancy-related risk factors over the life course.^7^ HDP may also trigger or accelerate vascular ageing,^8^ which could also contribute to existing pathological processes, or may itself be a driving mechanism for increased CVD risk.^4^

Most published studies assessing CVD risk in people with a history of HDP have focused on non-arrhythmic cardiovascular outcomes (e.g., myocardial infarction, stroke, heart failure, and coronary artery disease).^7, 9, 10^ However, the global prevalence of atrial fibrillation (AFib) has rapidly increased over the past three decades,^11^ rising from 19 million individuals in 1990 to 38 million in 2017 (100% increase).^11^ Indeed, AFib is now considered the cardiovascular epidemic of the 21^st^ century,^12^ with the absolute burden of the disease projected to rise by more than 60% by 2025,^12^ underscoring the critical need to study AFib to better understand its unique risk factors.

Prior studies examining the association between HDP and incident AFib^4, 13–16^ have typically considered AFib as part of a composite outcome^13, 15^ or have focused solely on preeclampsia.^15, 17, 18^ Further, most of these studies had relatively short (median) follow-up times (e.g., one,^14^ two,^17^ and seven^19^ years), did not consider death as a competing risk, and failed to control for several important confounders. Further, despite evidence that HDPs may represent distinct disease phenotypes,^7, 20^ which present with varying levels of acute severity, no studies have examined potential dose-response associations between distinct HDP subtypes [e.g., chronic hypertension in pregnancy, *(de novo)* gestational hypertension] and incident AFib.

Given the rising social, economic, and public health burden of AFib,^11, 21^ and a growing body of evidence suggesting that females are more likely to be symptomatic,^22^ report lower overall quality of life, experience greater functional impairment,^23^ have a higher susceptibility to AFib as their blood pressure increases,^24^ and are diagnosed later in the disease course compared to males,^25^ more evidence is needed about the relationship between HDP and its subtypes as sex-specific risk factors for AFib.^14^ To address these knowledge gaps, our *primary aim* was to examine the association between exposure to any HDP in a first singleton pregnancy and subsequent incident AFib using a competing risks analytic framework. Our *secondary aim* was to evaluate potential dose-response relationships between the severity of HDP (based on specific HDP subtypes) and incident AFib using the same analytical approach.

## Methods

This study followed published methodological guidance on assessing CVD risk in people with a history of HDP using administrative healthcare data.^7^ We also used the REporting of studies Conducted using Observational Routinely-collected health Data (RECORD) statement^26^ to guide study reporting. A copy of our completed RECORD checklist is provided in Table S1.

We acknowledge that both gender expression and biological sex exist on a spectrum and have striven to use gender neutral terminology as much as possible. When the terms women/woman and maternal are used this manuscript, they should be interpreted inclusively.

### Data Availability Statement

The dataset used in this study is held securely in coded form at ICES. While legal data sharing agreements between ICES and data providers (e.g., healthcare organizations and government) prohibit ICES from making the dataset publicly available, access may be granted to those who meet pre-specified criteria for confidential access, available at www.ices.on.ca/DAS. The full dataset creation plan and underlying analytic code are available from the authors upon request, understanding that the computer programs may rely upon coding templates or macros that are unique to ICES and are therefore either inaccessible or may require modification.

### Study Design and Setting

This was a population-based retrospective cohort study of nulliparous women discharged from Ontario hospitals for obstetrical delivery of their first live or stillborn singleton infant between April 1, 2002 and December 31, 2017 (index date of cohort entry). Subjects were followed until they had an incident AFib diagnosis, a recorded death date, were no longer eligible for OHIP (i.e., migrated out of Ontario), or at the end of study follow-up (December 31^st^, 2019), whichever came first.

Ontario is Canada’s most populous province and provides essential medical services, in-hospital prescription drugs, and immunizations free of charge for all residents through the Ontario Health Insurance Program (OHIP). Data generated from the use of routine healthcare services are maintained at ICES. ICES is an independent, non-profit research institute whose legal status under Ontario’s health information privacy law allows it to collect and analyze health care and demographic data, without consent, for health system evaluation and improvement.

### Data Sources

We used data obtained from record-level, coded, population-based administrative databases that were linked using unique encoded identifiers and analyzed at ICES (Figure S1; details about each dataset are provided in Table S2). Briefly, demographic information at index delivery (maternal date of birth, residential postal code) was obtained from the Registered Persons Database (RPDB), a population registry file of all people eligible for OHIP since 1991 and enables linkage with other data holdings at ICES. The mother-baby database (MOMBABY), which uses the Canadian Institutes for Health Information Discharge Abstract Database (CIHI-DAD) to link in-hospital maternal and infant delivery records, was used to obtain information about maternal parity as well as baby’s sex, date of birth, birthweight, and gestational age at birth. Information about diagnoses and procedures from clinical encounters and were obtained from the National Ambulatory Care Reporting System (NACRS), OHIP, CIHI-DAD, and the Same Day Surgery (SDS) database, as well as several ICES-derived cohorts (Ontario Hypertension Dataset, Ontario Diabetes Dataset, Ontario Crohn’s and Colitis Cohort, Ontario Rheumatoid Arthritis Dataset).

We used the Ontario portion of the Immigration, Refugee and Citizenship Canada’s (IRCC) permanent resident database to ascertain immigration status. Information about area-level socioeconomic status was ascertained using the Ontario Marginalization Index (ON-Marg; quintiles)^27^ by linking residential postal codes (RPDB) with census data, and we used the Rural Index of Ontario (RIO) to determine subject rurality at index delivery. The postal code conversion file was used to convert subject postal codes to area-level quintiles.

### Subject Eligibility

All adults aged ≥18 years who delivered their first live or stillborn infant at ≥20 weeks gestational age, or >500-gram birthweight, between April 1, 2002 and December 31, 2017 (index date) were initially eligible for inclusion. Individuals were excluded if they were >55 years of age, not Ontario residents, had an invalid health card number, a recorded death date prior to their index date, were not eligible for OHIP coverage, or had <2 years of continuous coverage, prior to cohort entry. People with a history of/ongoing AFib at the time of index delivery and those missing data required for ON-Marg (n= 8,587; 1.1%) were also excluded. During the exposure accrual period, approximately 98% of all births in Ontario occurred in-hospital.^28^

### Study Variables

A directed acyclic graph (DAG) illustrating the hypothesized relationships between study variables (i.e., primary exposure, outcome, relevant confounders) is shown in Figure S2.

#### Primary and Secondary Exposures

The primary exposure was a composite ‘any HDP’ variable defined as any diagnosis of gestational hypertension, chronic hypertension in pregnancy, preeclampsia, chronic hypertension with superimposed preeclampsia (superimposed preeclampsia), eclampsia, or unspecified HDP in subjects’ first obstetrical delivery. Secondary exposures included each individual HDP within the composite HDP exposure.

No case-finding definitions/algorithms for ‘any HDP’, or individual HDP subtypes, have been validated on Ontario population-based administrative data. As such, each HDP exposure was identified by applying the best available, and most relevant, case-finding definition validated in other jurisdictions to each subject’s discharge abstract (Table S3).^7, 29^ We also searched for ‘unspecified’ HDP by looking for any International Classification of Disease, Tenth Revision (ICD-10) code beginning with O16 in each subject’s obstetrical discharge abstract.

We employed a hierarchical ‘severity coding’ approach to exposure classification whereby any subject whose discharge abstract met the case-finding definition for >1 HDP subtype had their exposure status re-coded such that only their most severe diagnosis was recorded. Further details on the use of this approach are provided in Data Supplement S4.

#### Outcome and Competing Risk

The primary outcome was incident AFib, which was identified using a case-finding definition previously validated in Ontario population-based administrative data (Table S4).^30^ This algorithm has a reported specificity of 99%, and a positive predictive value of 71%, for the identification of incident AFib in Ontario adults ≥20 years of age.^30^ We did not differentiate between AFib type (i.e., paroxysmal, persistent, long-standing). The competing risk, all-cause death, was determined by the presence of a death date in the RPDB.

#### Confounders

We identified several risk factors for AFib that are also associated with, but not an effect of, HDP (*i.e.,* confounders)^31^ based on input from clinical experts and evidence available from published literature.^32–38^ Behavioural and clinical factors identified as potential confounders included: maternal age at delivery, pre-existing diabetes mellitus, polycystic ovary syndrome, sleep apnea, smoking, kidney disease, chronic immune-mediated inflammatory conditions, and pre-pregnancy obesity. We used ON-Marg material resources dimension^27^ quintiles to control for potential confounding by area-level socioeconomic status.^39, 40^ All confounders were treated as time-fixed and assessed only at baseline. Further details about the methods used to identify these conditions are summarized in Table S4.

#### Follow-up for Outcome and Competing Risk

Subjects were followed until they had an incident AFib diagnosis (outcome), a recorded death date in the RPDB (competing risk), were no longer eligible for OHIP, or at the end of study follow-up, whichever came first. All subjects who were eligible for OHIP, were still alive, and did not fulfil the algorithm for AFib by December 31^st^, 2019 were censored.

### Statistical Methods

#### Descriptive Analyses

Baseline characteristics and comorbidities (i.e., conditions present at any time prior to index delivery) are presented as medians (quartile (Q)3 – Q1) and number affected (n), percentages (%) for categorical variables. We assessed differences in baseline characteristics between HDP exposure status using absolute standardized differences (ASD). ASD >10% were considered indicative of potentially meaningful differences between the exposure and characteristic being assessed.^41^ We also calculated the proportion of subjects diagnosed with each HDP subtype over the study period. In accordance with ICES privacy policies, small cells (counts <6) have been suppressed or combined to reduce the risk of reidentification.

#### Crude and Multivariable Cause-Specific Cox Proportional Hazards Regression

Prior to fitting statistical models, each study variable was assessed for missingness, the presence of extreme/influential values, and possibly erroneous data (as appropriate). Phi coefficients were used to assess the strength of correlation between dichotomous variables; coefficients of ≥0.2 were interpreted as at least moderately correlated variables.^42^

#### Cause-Specific Cox Proportional Hazards Regression

We used competing-risks Cox proportional hazards regression to compute crude and multivariable-adjusted cause-specific hazard ratios (csHRs), and accompanying 95% CIs, to assess the association between history of any HDP, and specific HDP subtypes and (a) incident AFib in those still alive during follow-up as well as (b) all-cause mortality in the absence of a prior AFib diagnosis.

We fit separate models for incident AFib and all-cause mortality, treating subjects who died, or were diagnosed with AFib, as censored in each respective model. We also tested for multiplicative interaction between HDP and material resources given known disparities in the burden of HDP and AFib according to socioeconomic status.^43, 44^ Calendar time was used as the underlying time scale in all models.

For each cause-specific model, we used both graphical and formal statistical tests (Schoenfeld residuals and residual plots,^45^ Kolmogorov-type supremum tests^46^) to assess whether the effect of each variable on the csHazard function was constant over time.^47^ A 2-sided p-value <0.05 was considered evidence that the variable being tested likely violated the PH assumption. ^46, 48^

After visually assessing residual plots, and considering the results of formal statistical tests, the PH assumption was considered violated for gestational hypertension (AFib model only), as well as the covariates age (both cause-specific models) and material resources (mortality models only). As such, in AFib models for subjects exposed to gestational hypertension, we performed a time-segmented analysis by restricting follow-up time to >4.5 years post-delivery, as the PH assumption held for this follow-up period. In models assessing all-cause mortality, we stratified by material resources.^49^ Final models included maternal age at delivery as a restricted cubic spline with five knots placed at the 5^th^, 27.5^th^, 50^th^, 72.5^th^, and 95^th^ percentiles.^50^

#### Sensitivity Analyses

We carried out three sensitivity analyses: two were carried out to assess how (1) excluding subjects with certain pre-existing conditions (Table S5), and (2) restricting follow-up times for observing AFib/mortality events, impacted effect estimates. A third sensitivity analysis assessed the potential impact of unmeasured confounding on the effect estimates generated (E-values). Further details are provided in Data Supplement S6.

All analyses were carried out using SAS Enterprise Guide (Version 8.3, SAS Institute, Cary, NC). Forest plots were generated using the ggplot2 package^51^ in RStudio (v. 2023.12.1.402) to visualize effect estimates (95% CIs) and facilitate comparisons between HDP exposures.

### Ethics Approval

The use of the data in this project is authorized under section 45 of Ontario’s Personal Health Information Protection Act (PHIPA) and does not require review by a Research Ethics Board.

## Results

### Selection of Study Subjects

We identified 2,105,409 women discharged from an Ontario hospital for a potentially eligible obstetrical delivery between April 1, 2002 and December 31, 2017. After excluding non-Ontario residents, people <18 and >55 years of age, individuals with a recorded death date prior to their index date, <2 years of continuous OHIP eligibility, and a history of/ongoing AFib prior to or at index delivery, in addition to non-nulliparous women and subsequent deliveries of otherwise eligible women, subjects with multiple gestation first pregnancies, and people with missing ON-Marg information, a total of 771,521 subjects were included in the study cohort (Figure 1).

**Figure 1.**
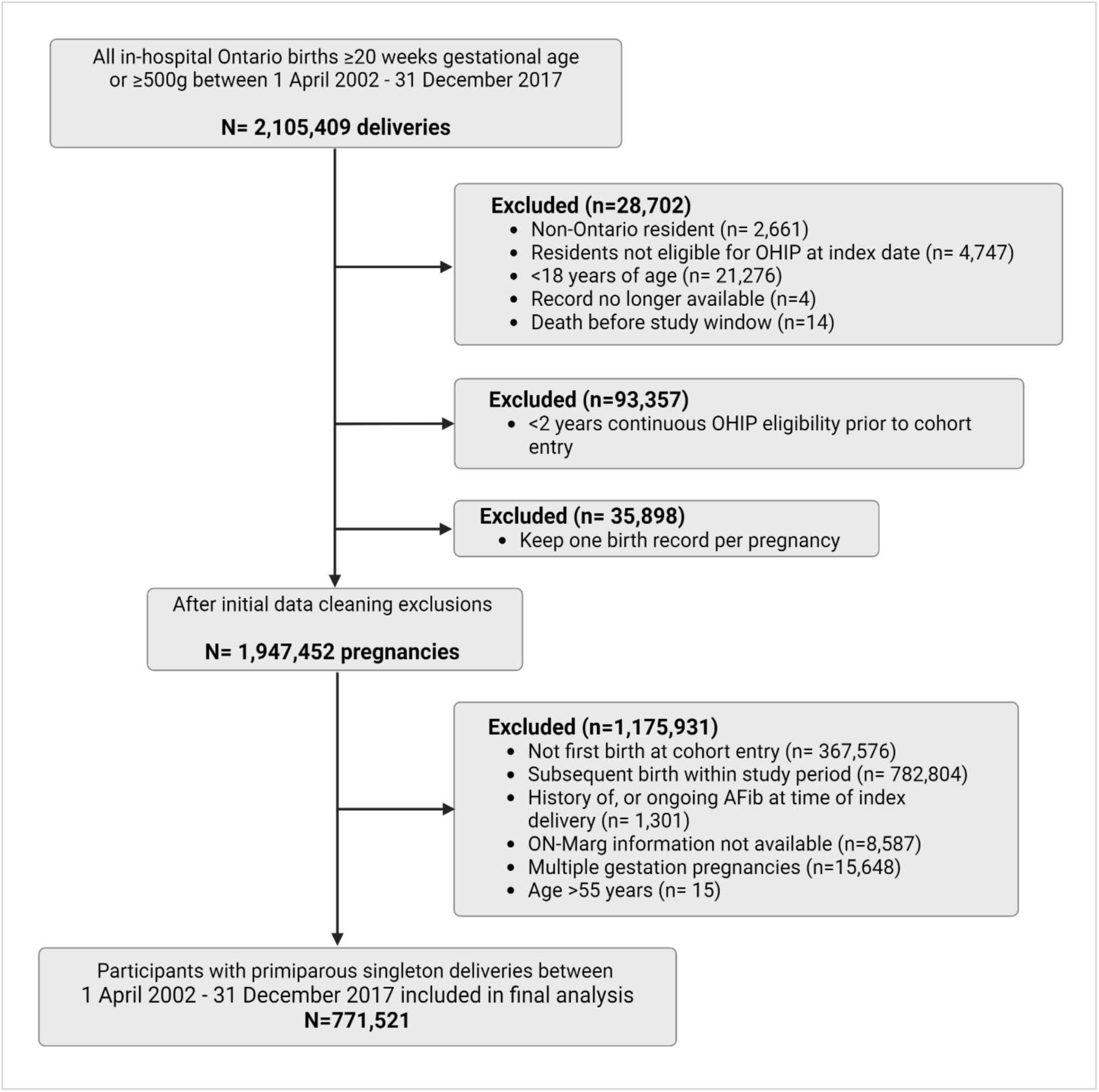
Flow diagram illustrating the selection of study subjects.

### Baseline Maternal Characteristics at Study Entry

Most maternal risk factors and adverse pregnancy outcomes were more prevalent in people with a history of HDP. However, only a moderate number of differences were considered potentially meaningful (ASD >10%); Table 1.

**Table 1.** Baseline maternal characteristics at study entry.

|  | Total | No HDP | HDP | ASD (%) |
| --- | --- | --- | --- | --- |
| Total No. Study Subjects (%) | 771,521 (100) | 710,035 (92.0) | 61,486 (8.0) | -- |
| **Maternal Characteristics** |  |  |  |  |
| **Age at First Delivery, Years** |  |  |  |  |
| Median (Q1-Q3) | 29 (25-32) | 29 (25-32) | 29 (26-33) | **13** |
| **Age Group (Years)** |  |  |  |  |
| <20 | 34,697 (4.5) | 32,613 (4.6) | 2,084 (3.4) | 5 |
| 20 to 24 | 132,764 (17.2) | 123,448 (17.4) | 9,316 (15.2) | 2 |
| 25 to 29 | 250,841 (32.5) | 231,439 (32.6) | 19,402 (31.6) | 6 |
| 30 to 34 | 244,567 (31.7) | 225,397 (31.7) | 19,170 (31.2) | 6 |
| 35 to 39 | 90,178 (11.7) | 81,204 (11.4) | 8,974 (14.6) | 2 |
| 40 to 44 | 17,258 (2.2) | 14,976 (2.1) | 2,282 (3.7) | 1 |
| 45 to 49 | 1,117 (0.1) | 889 (0.1) | 228 (0.4) | 9 |
| 50+ | 99 (0.01) | 69 (0.01) | 30 (0.05) | **10** |
| **Comorbid Conditions\*** |  |  |  |  |
| Diabetes Mellitus | 11,296 (1.5) | 8,828 (1.2) | 2,468 (4.0) | **17** |
| Polycystic Ovary Syndrome | 26,192 (3.4) | 23,229 (3.3) | 2,963 (4.8) | 8 |
| Sleep Apnea** | 15,148 (2.0) | 13,102 (1.8) | 2,046 (3.3) | 9 |
| Ever Smoker | 23,730 (3.1) | 21,676 (3.1) | 2,054 (3.3) | 2 |
| Kidney Disease | 18,337 (2.4) | 16,060 (2.3) | 2,277 (3.7) | 9 |
| Chronic Immune-Mediated Inflammatory Conditions† | 16,569 (2.1) | 14,942 (2.1) | 1,627 (2.6) | 4 |
| Obesity | 2,359 (0.3) | 1,788 (0.3) | 571 (0.9%) | 9 |
| Baseline Hypertension | 16,715 (2.2) | 9,737 (1.4) | 6,978 (11.3%) | **42** |
| **Sociodemographic Characteristics** |  |  |  |  |
| Immigrant†† | 159,375 (20.7) | 149,546 (21.1%) | 9,829 (16.0%) | **13** |
| **Rurality Index of Ontario (RIO2008)\*** |  |  |  |  |
| Urban | 579,948 (75.2) | 536,410 (75.5) | 43,538 (70.8) | **11** |
| Suburban | 140,165 (18.2) | 126,912 (17.9) | 13,253 (21.6) | 9 |
| Rural | 47,714 (6.2) | 43,371 (6.1) | 4,343 (7.1) | 4 |
| Missing | 3,694 (0.5) | 3,342 (0.5) | 352 (0.6) | 1 |

| **ON-Marg Dimension** |  |  |  |  |
| --- | --- | --- | --- | --- |
| **Age and Labour Force Dimension** |  |  |  |  |
| Quintile 1 | 250,642 (32.5) | 231,223 (32.6) | 19,419 (31.6) | 2 |
| Quintile 5 | 103,732 (13.4) | 94,977 (13.4) | 8,755 (14.2) | 3 |
| **Material Resources Dimension** |  |  |  |  |
| Quintile 1 | 166,892 (21.6) | 153,917 (21.7) | 12,975 (21.1) | 1 |
| Quintile 5 | 158,854 (20.6) | 145,886 (20.5) | 12,968 (21.1) | 1 |
| **Households and Dwellings Dimension** |  |  |  |  |
| Quintile 1 | 149,548 (19.4) | 138,366 (19.5) | 11,182 (18.2) | 3 |
| Quintile 5 | 184,498 (23.9) | 170,461 (24.0) | 14,037 (22.8) | 3 |
| **Racialized and Newcomer Populations Dimension** |  |  |  |  |
| Quintile 1 | 106,365 (13.8) | 96,557 (13.6) | 9,808 (16.0) | 7 |
| Quintile 5 | 234,329 (30.4) | 218,336 (30.8) | 15,993 (26.0) | **11** |
| **Visible Minority Quintile** |  |  |  |  |
| Quintile 1 | 161,174 (20.9) | 146,182 (20.6) | 14,992 (24.4) | 9 |
| Quintile 5 | 125,948 (16.3) | 117,881 (16.6) | 8,067 (13.1) | 10 |
| Missing | 3,960 (0.5) | 3,684 (0.5) | 276 (0.4) | 1 |

Note: Values are n (%) unless otherwise noted. An ASD >10% (**bold**) was interpreted as a potentially meaningful difference in a baseline characteristic between groups.
ASD= absolute standardized difference, SD= standard deviation, ON-Marg = Ontario Marginalization Index, OHIP = Ontario Health Insurance Program, IRCC = Immigration, Refugee, and Citizenship Canada, Q= quartile
<sup>†</sup> Composite variable that includes subjects diagnosed with ≥1 of the following conditions prior to cohort entry: lupus, inflammatory bowel disease, psoriatic arthritis, ankylosing spondylitis, and/or rheumatoid arthritis.
<sup>\*</sup>RIO2008 categorized as Urban = RIO < 10, Suburban = RIO 10–39, Rural = RIO ≥ 40
<sup>\*\*</sup>Proxy variable based on having ≥1 OHIP code for sleep study.
<sup>††</sup> Has an IRCC landing date

Individuals with a history of HDP in their first obstetrical delivery were older than those with no HDP diagnosis (ASD for median age, 13%) and were more likely to have pre-existing diabetes and hypertension. In contrast, subjects with a history of HDP were less likely to be an immigrant, to reside in an urban area, or in neighborhoods with the highest quintile of racialized and newcomer populations (Table 1). The prevalence of pre-existing cardiovascular and endocrine conditions at baseline was numerically equivalent to, or marginally higher in, subjects diagnosed with HDP (Table S6). Additional details related to baseline obstetrical characteristics are provided in Table S7.

### Hypertensive Disorder of Pregnancy Exposure Status and Incident Chronic Hypertension Post-delivery

Approximately 8% of study subjects were diagnosed with HDP during the 16-year exposure accrual period (79.7 per 1,000 deliveries, n=61,486). The most common diagnosis was gestational hypertension (52.4 per 1,000 deliveries, n=40,460); the least common was eclampsia (1.1 per 1,000 deliveries, n=824). A total of 1,691 subjects had a diagnostic code for unspecified HDP (2.2 per 1,000 deliveries) (Figure 2).

**Figure 2.**
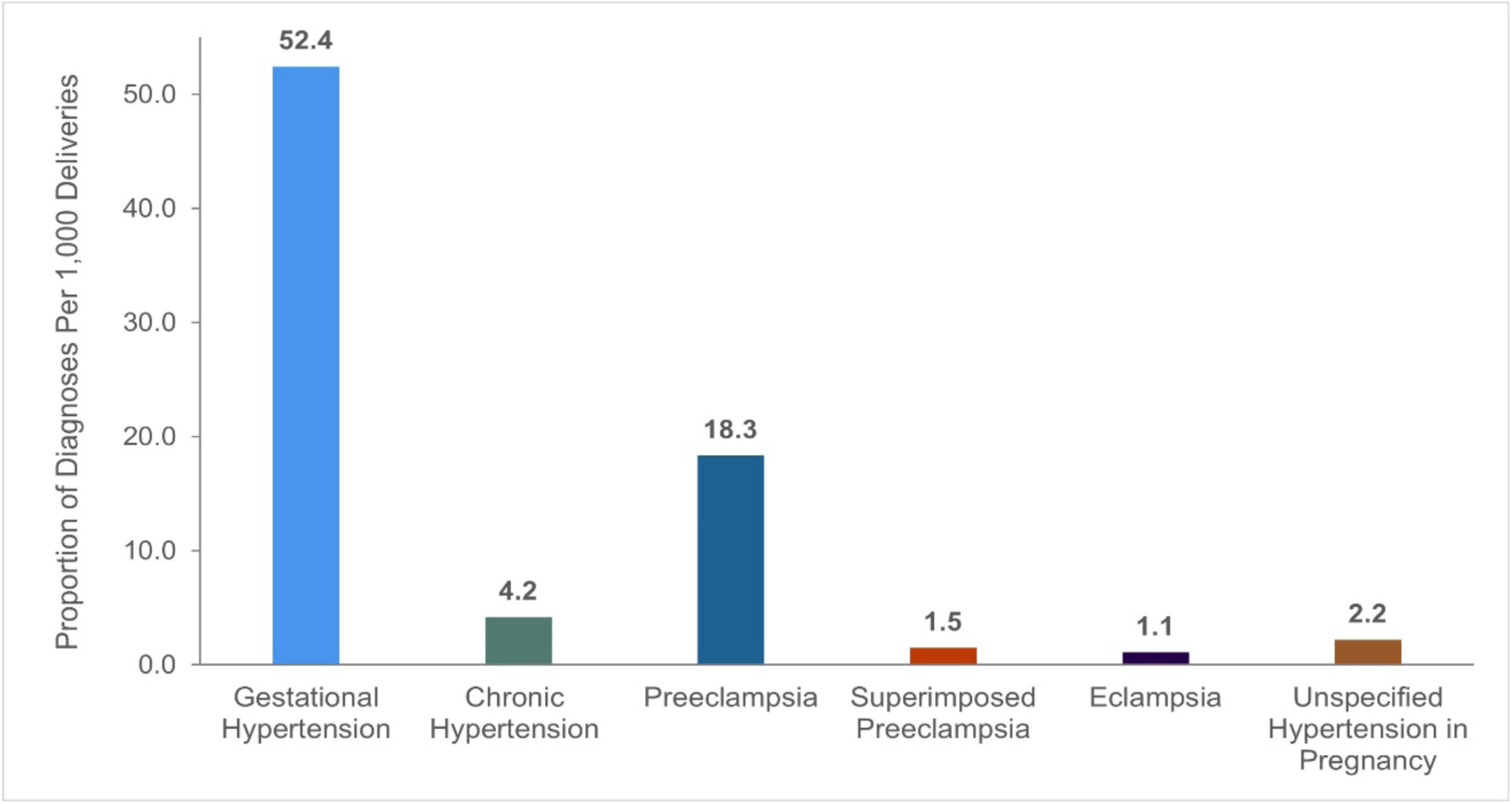
Proportion of study subjects (per 1,000 deliveries) diagnosed with a hypertensive disorder of pregnancy during the study’s exposure accrual period, by subtype. *For each HDP subtype, the numerator included the total number of subjects diagnosed with that specific subtype during the study period and the denominator included the total number of study subjects*.

Nearly 5% (n= 36,507) of study subjects were diagnosed with incident chronic hypertension at some point after pregnancy during the follow-up period. It was especially prevalent in subjects diagnosed with gestational hypertension (n=6,486, 16%) and preeclampsia (n=2,030, 14%) in their index delivery.

### Incidence Rates and Time to Event Analyses

The total person-time of follow-up was 7,380,304 person-years (median follow-up 9.5 years), during which there were 2,483 (0.3%) incident AFib diagnoses and 2,951 (0.4%) deaths. The crude cause-specific incidence rate (95% CI) of AFib ranged from 4.52 (3.96- 5.08) per 10,000 person-years for gestational hypertension to 10.18 (7.13-13.24) per 10,000 person-years for chronic hypertension in pregnancy. Crude mortality rates (95% CI) in those without a prior AFib diagnosis ranged from 4.59 (4.03-5.16) per 10,000 person-years for gestational hypertension to 11.97 (6.29-17.66) per 10,000 person-years for superimposed preeclampsia; Table 2.

**Table 2.** Total number of events, person-years of follow-up, cause-specific incidence rates, and incidence rate ratios for incident atrial fibrillation and all-cause mortality without a prior atrial fibrillation diagnosis by any hypertensive disorder of pregnancy and individual subtypes.

|  | Exposure |  |  |  |  |  |  |  |
| --- | --- | --- | --- | --- | --- | --- | --- | --- |
|  | Not Exposed | Any HDP | Gestational Hypertension | Chronic Hypertension | Preeclampsia | Superimposed Preeclampsia | Eclampsia | Unspecified |
| **Incident Atrial Fibrillation** |  |  |  |  |  |  |  |  |
| Total events | 2,187 | <300* | 178 | 30 | 65 | <10* | <10* | 10 |
| Total PY of follow-up | 6,797,092 | 583,212 | 393,969 | 29,456 | 123,329 | 10,022 | 9,207 | 17,229 |
| csIR per 10,000 PY (95% CI) | 3.2 (3.1-3.3) | 5.1 (4.6-5.6) | 4.5 (4.0 - 5.1) | 10.2 (7.1-13.2) | 5.3 (4.2-6.4) | * | * | 5.8 (2.8-8.8) |
| csIRR (95% CI) | **Reference** | 1.58 (1.40-1.78) | 1.40 (1.21-1.64) | 3.17 (2.21-4.54) | 1.64 (1.28-2.10) | * | * | 1.80 (0.97-3.36) |
| **All-cause Mortality without a Prior Atrial Fibrillation Diagnosis** |  |  |  |  |  |  |  |  |
| Total events | 2,631 | 320 | 181 | 32 | 73 | 12 | 8 | 14 |
| Total PY of follow-up | 6,797,092 | 583,212 | 393,969 | 29,456 | 123,329 | 10,022 | 9,207 | 17,229 |
| csIR per 10,000 PY (95% CI) | 3.9 (3.8-4.0) | 5.5 (5.0-6.0) | 4.6 (4.0-5.2) | 10.9 (7.7-14.0) | 5.9 (4.8-7.1) | 12.0 (6.3-17.7) | 8.7 (3.6-13.7) | 8.1 (4.6-11.7) |
| csIRR (95% CI) | **Reference** | 1.42 (1.26-1.59) | 1.19 (1.02-1.38) | 2.81 (1.98-3.98) | 1.53 (1.21-1.93) | 3.09 (1.75-5.45) | 2.24 (1.12-4.49) | 2.10 (1.24-3.55) |

Abbreviations: PY = person-years, CS= cause-specific, CI = confidence interval, IR = incidence rate, HR = hazard ratio
All calculations made using the fmsb package (v.0.7.6)<sup>73</sup> in RStudio (v. 2023.12.1.402).
\* Data suppressed due to small numbers to comply with ICES privacy requirements

The median age (Q1-Q3) of subjects with AFib was 30 (26-34 years), with 32% aged 40 years or older at their time of diagnosis, and the median age of those who died was 29 years (23-33 years), with 33% aged 40 years of age or older at their time of death. A total of 766,087 subjects were censored [n=26,926 (3.5%) due to a lapse in OHIP coverage; n= 739,161 (95.8%) at end of study].

When HDP exposure status was considered collectively (i.e., exposure to any HDP subtype), the median time to event was nearly identical for both AFib and all-cause mortality [7.1 years (2,574 days)] vs. [2,567 days (7.0 years)], respectively). However, when each HDP exposure was considered individually, there was considerable heterogeneity with respect to median time-to-event for both event types (Figure 3). Specifically, subjects diagnosed with preeclampsia had the shortest median time to an incident AFib diagnosis at approximately 5.5 years, while those diagnosed with eclampsia had the longest median time to an AFib diagnosis at approximately 11.8 years. Further, the median time to an incident AFib diagnosis was most similar between subjects diagnosed with unspecified HDP and chronic hypertension in pregnancy (6.1 and 6.4 years, respectively), while the median time to death was most similar between those diagnosed with unspecified HDP and superimposed preeclampsia (8.7 and 8.3 years, respectively).

**Figure 3.**
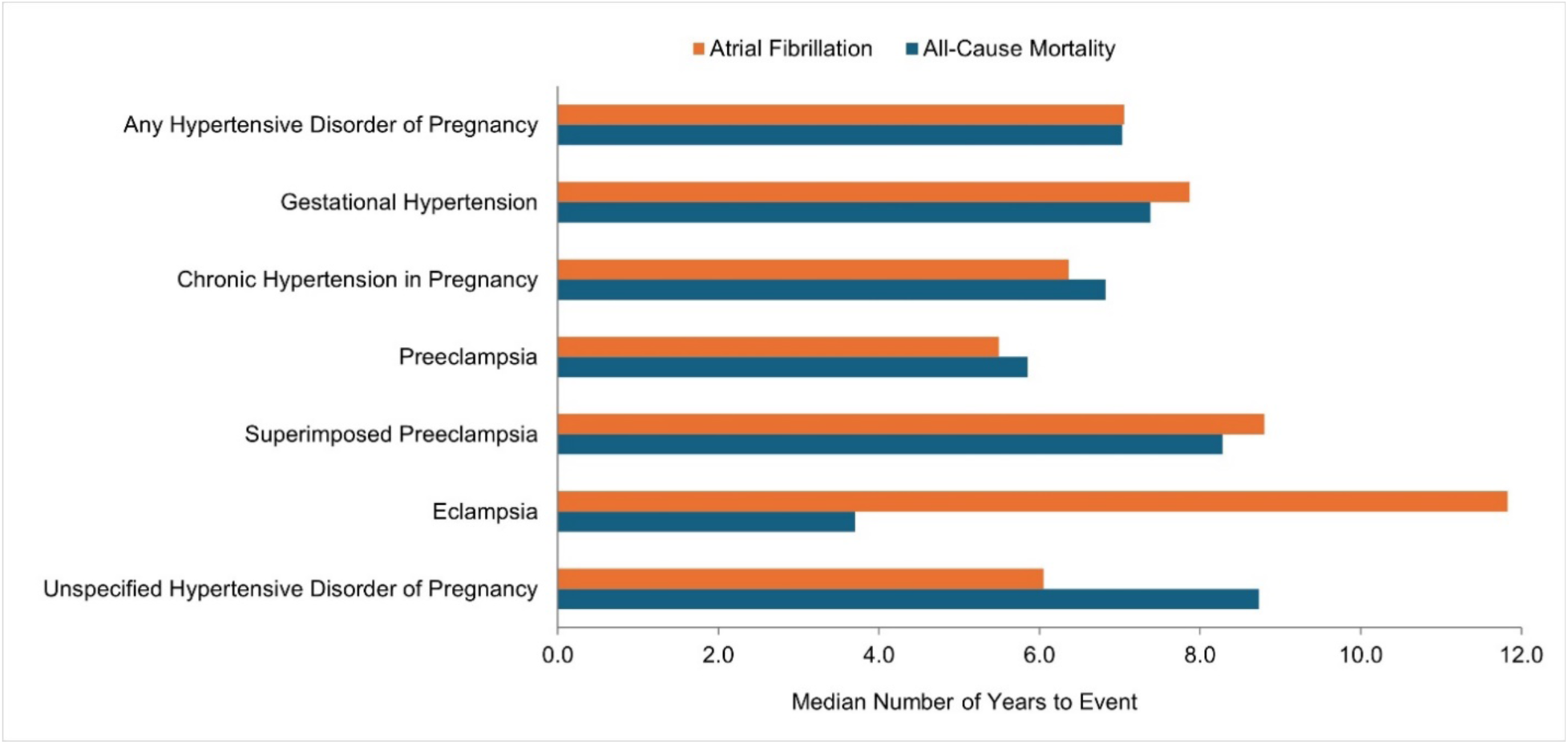
Median number of years to incident atrial fibrillation and all-cause mortality in study subjects diagnosed with a hypertensive disorder of pregnancy, overall and by exposure subtype. *Note: Results for eclampsia should be interpreted with caution due to the low number of events in subjects diagnosed with this HDP subtype (n <10)*.

### Cause-Specific Cumulative Incidence Curves

Cause-specific cumulative incidence curves for incident AFib and all-cause mortality, stratified by HDP exposure status, are shown in Figure 4. Subjects with a history of any HDP in their first singleton delivery had a consistently higher incidence of AFib and all-cause mortality compared to their unexposed counterparts over the follow-up period; differences in these curves were apparent within approximately two years of the pregnancy. For all-cause mortality, clear differences in these curves occurred slightly later, at approximately three years post-delivery (Figure 4). Similar findings were noted for each HDP subtype (Figures S4, S5).

**Figure 4.**
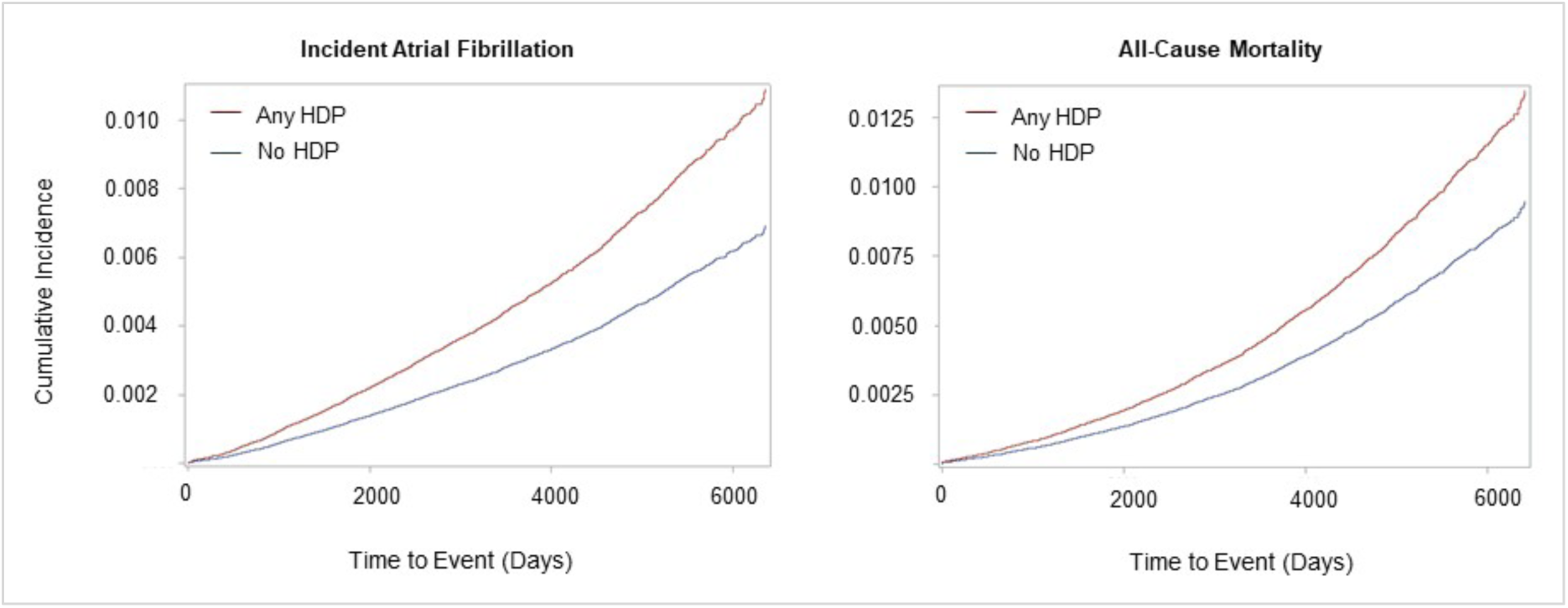
Cause-specific cumulative incidence curves for atrial fibrillation (outcome) and all-cause mortality (competing risk) stratified by hypertensive disorder of pregnancy exposure status (first obstetrical delivery).

### Cause-Specific Hazard of AFib and All-Cause Mortality

After adjusting for confounders, we found that a history of any HDP in a first obstetrical delivery was associated with a 45% increased csHazard of incident AFib (adjusted csHR 1.45, 95% CI: 1.28-1.64), as well as an increased csHazard of death in subjects without a prior AFib diagnosis (adjusted csHR 1.31, 95% CI: 1.16-1.47) (Figures 5A and 5B, respectively).

**Figure 5.**
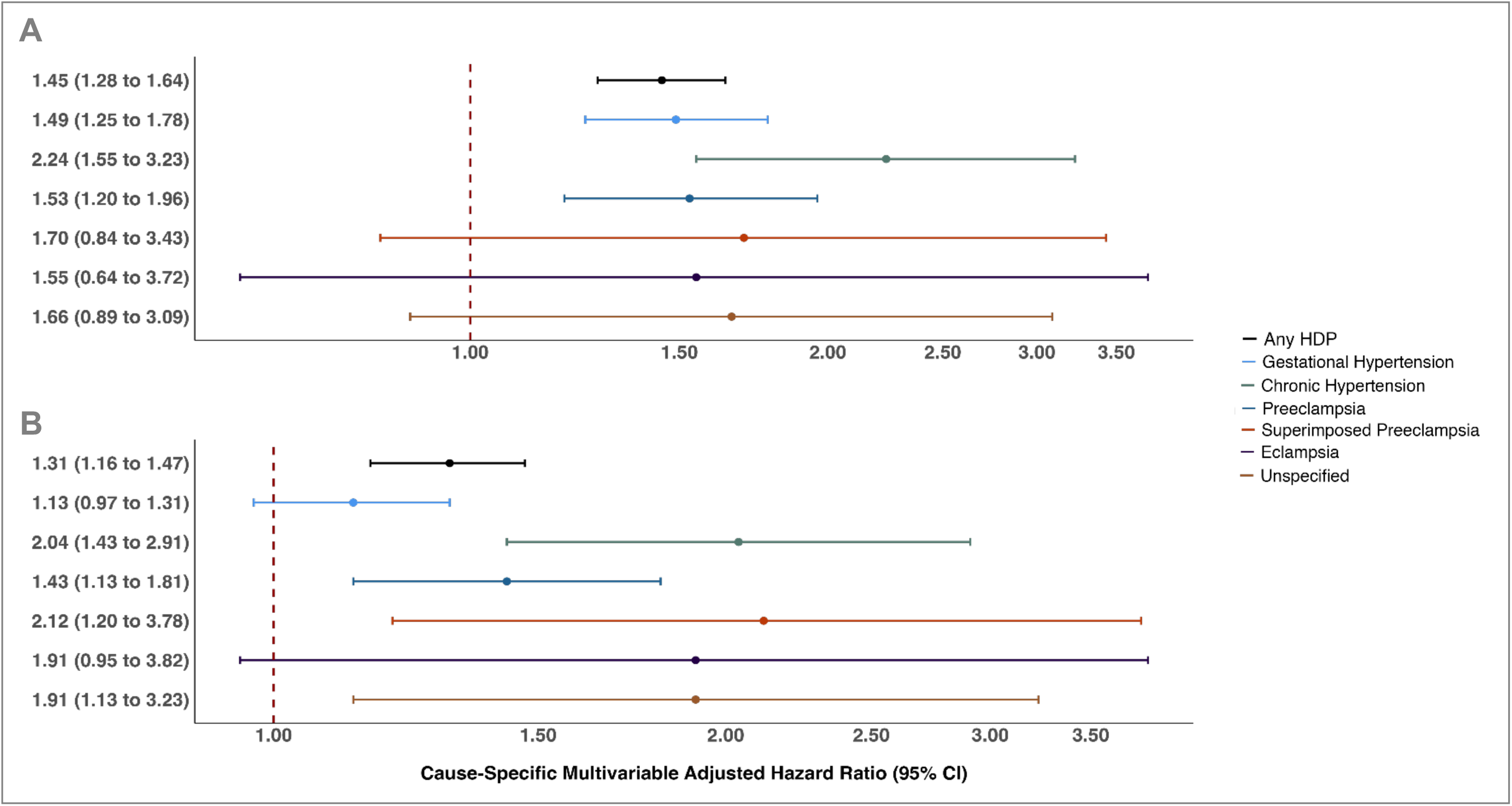
Cause-specific multivariable adjusted hazard ratios and accompanying 95% confidence intervals (log-scale), for atrial fibrillation (A) and all-cause mortality (B) by any hypertensive disorder of pregnancy and each subtype. *Note: Hazard ratios adjusted for maternal age at delivery, diabetes mellitus, polycystic ovary syndrome, sleep apnea, smoking, kidney disease, obesity, chronic immune-mediated conditions, and ON-Marg material resources dimension (quintiles)*.

With respect to specific HDP subtypes, we found that a history of gestational hypertension, preeclampsia, and chronic hypertension in pregnancy were all associated with an increased csHazard of incident AFib [adjusted csHR (95% CI): 1.49 (1.25-1.78), 1.53 (1.20-1.96), and 2.24 (1.55-3.23), respectively]; Figure 5A. While crude estimates indicated a significant cause-specific association between superimposed preeclampsia and incident AFib, the observed relationship was attenuated and no longer significant after multivariable adjustment (csHR 1.70, 95% CI: 0.84 to 3.43); Table S8. We also found that a history of preeclampsia, unspecified HDP, chronic hypertension in pregnancy, and superimposed preeclampsia were each independently associated with an increased csHazard of death in subjects without an AFib diagnosis [adjusted csHR (95% CI): 1.43 (1.13-1.81), 1.91 (1.13-3.23), 2.04 (1.43-2.91), and 2.12 (1.20-3.78), respectively]; Figure 5B. Crude estimates also indicated a significant cause-specific association between both gestational hypertension and eclampsia and all-cause mortality. However, these relationships were attenuated and no longer significant after multivariable adjustment [csHR (95% CI): 1.13 (0.97-1.31) and 1.91 (0.96-3.83), respectively]; Table S9.

### Dose-Response Relationships

As summarized in Table 3, we identified associations suggestive of ‘dose-response’ relationship whereby both the severity of *de novo* HDP diagnoses, and exposure to pre-pregnancy chronic hypertension, were associated with higher cause-specific rates of AFib and all-cause mortality. Specifically, a history of gestational hypertension in a first delivery showed the smallest significant cause-specific association with incident AFib [adjusted csHR (95% CI) 1.49 (1.25-1.78)] compared to more severe *de novo* HDP exposures, such as preeclampsia. However, a history of chronic hypertension in pregnancy was most strongly associated with incident AFib compared to all other HDP diagnoses, with an adjusted csHR (95% CI) of 2.24 (1.55-3.23). We also noted that chronic hypertension in pregnancy was more strongly associated with all-cause mortality without a prior AFib diagnosis compared to preeclampsia [csHR (95%CI) 2.04 (1.43-2.91) vs. 1.43 (1.13- 1.81), a *de novo* diagnosis. Further, a history of exposure to both pre-pregnancy hypertension and preeclampsia (i.e., superimposed preeclampsia) in a first pregnancy showed the strongest cause-specific association with mortality compared to all other HDP subtypes (csHR 2.12, 95%CI: 1.20-3.78).

**Table 3.**
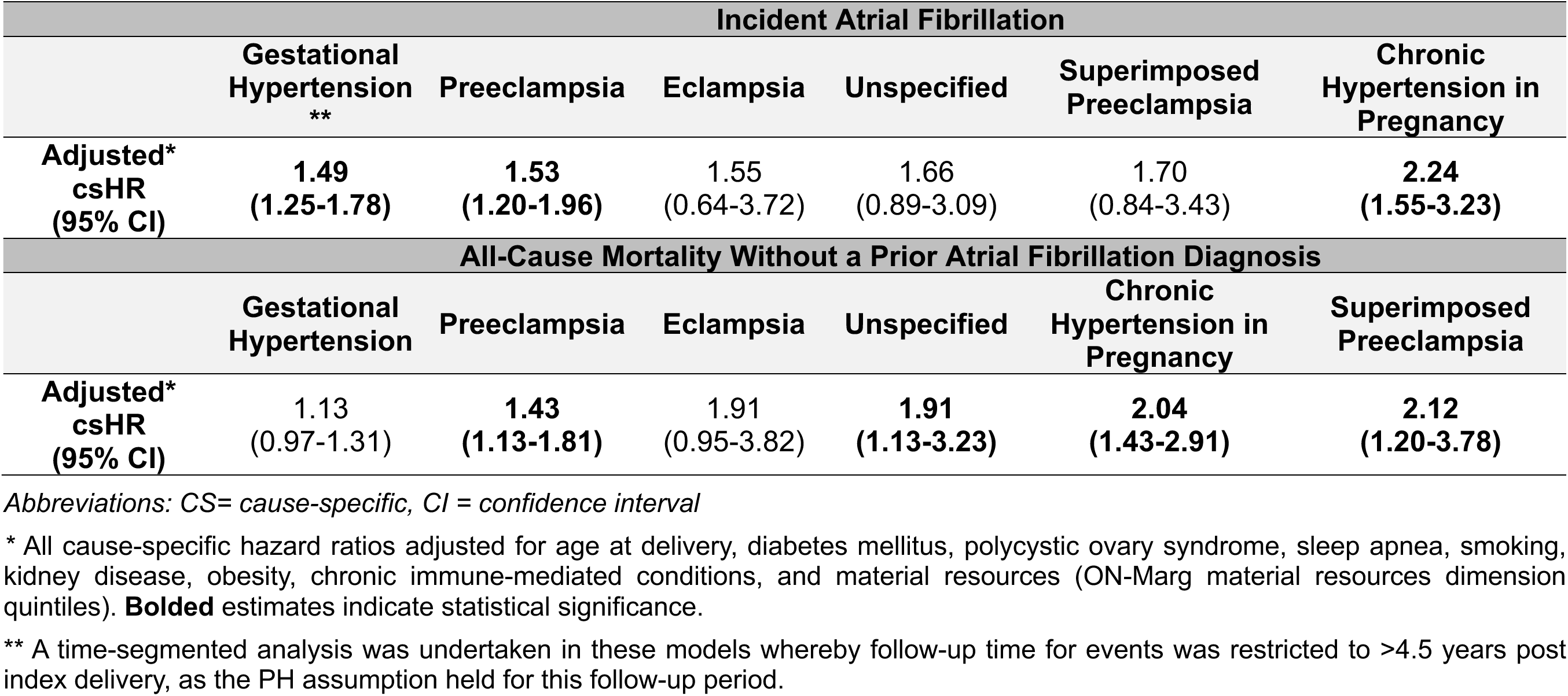
Cause-specific multivariable adjusted hazard ratios (95% confidence intervals) for incident atrial fibrillation and all- cause mortality without a prior atrial fibrillation diagnosis.

### Sensitivity Analyses

When the observation period for incident AFib events and all-cause mortality was limited to >90 days post index delivery, the csHazard of AFib in subjects who were still alive during follow-up was attenuated slightly in those exposed to any HDP, chronic hypertension in pregnancy, and preeclampsia in their first obstetrical delivery. However, all estimates remained statistically significant. Similar results were found for estimates of death without a prior AFib diagnosis (Table S10). Estimates obtained for models limited to >120 days of follow-up were similar to those limited to >90 days of follow up (not reported).

When models were re-run after excluding subjects with baseline cardiovascular and endocrine conditions, most estimates remained essentially unchanged or were minimally attenuated (Table S10). The only exception was the estimate for subjects exposed to chronic hypertension in pregnancy, where the csHR for AFib increased slightly from 2.24 (95% CI: 1.55-3.23) to 2.42 (95% CI: 1.63-3.59) after excluding subjects with these baseline conditions. Similar results were found for all-cause mortality without a prior AFib diagnosis after subjects with these pre-existing conditions were excluded from the analysis [adjusted csHR (95% CI): 2.05 (1.43-2.92) vs. 2.20 (1.51-3.19)].

E-values indicated that effect estimates were generally robust to unmeasured confounding (Table S11). Specifically, for gestational hypertension, chronic hypertension in pregnancy, and preeclampsia, an unmeasured confounder would need to at least double the risk of AFib and be at least twice as prevalent in those with a history of these HDPs, to explain away the observed associations. Similarly, for chronic hypertension in pregnancy, preeclampsia, superimposed preeclampsia, and unspecified HDP, an unmeasured confounder would need to at least double the risk of mortality before a prior AFib diagnosis and be at least twice as prevalent in those with a history of these exposures, to explain away the observed associations.

## Discussion

This population-based retrospective cohort study of 771,521 nulliparous women has three novel findings. First, a history of ‘any’ HDP is independently associated with an increased csHazard of both AFib and all-cause mortality, and these associations were observed in relatively young women (median time-to-event: 7 years postpartum). Second, a history of gestational hypertension, preeclampsia, and chronic hypertension in pregnancy are each independently associated with an increased csHazard of incident AFib, whereas a history of preeclampsia, unspecified HDP, chronic hypertension in pregnancy, and superimposed preeclampsia are each independently associated with an increased csHazard of all-cause mortality without a prior AFib diagnosis. Third, a dose-response relationship may exist within these associations, as both HDP severity, as well as the presence of pre-pregnancy chronic hypertension, were found to be associated with higher rates of both adverse outcomes.

The pathophysiologic mechanism(s) underlying the observed associations are complex and multifactorial.^7^ However, HDP and AFib share a number of important features,^52^ such as cardiac remodelling,^53^ endothelial dysfunction,^54, 55^ and inflammation,^56, 57^ which are likely central to the development of AFib in individuals with a history of HDP. This is further supported by evidence indicating that HDP–associated inflammation and endothelial dysfunction can persist well into the postpartum period,^52, 54, 56^ or become permanent,^58,59^ resulting in extensive changes to endothelial cell function in both small and large vessels.^60, 61^ Indeed, recent work^8^ also suggests that cardiovascular alterations associated with HDP lead to early vascular ageing, which could explain the increased predisposition to AFib noted in individuals exposed to HDP in this study– especially given that AFib is typically associated with older age.

Only a limited number of cohort studies have reported significant associations between a history of HDP and incident AFib.^4, 14, 16^ With respect to any HDP, we found a comparatively lower rate of incident AFib than that reported by Leon *et al.*,^16^ who reported a nearly 2-fold higher adjusted hazard of AFib in people exposed to HDP after 9.3 median years of follow-up. In comparison, after 9.5 median years of follow-up, we found a 1.4- fold higher adjusted csHazard of AFib, and a 1.3-fold higher adjusted csHazard of all-cause mortality in those with a history of any HDP. A few factors could explain these findings.

First, although Leon *et al.*^16^ controlled for some important confounders, such as maternal age and diabetes, several others (*e.g.,* pre-pregnancy body mass index (BMI), tobacco use, kidney disease) were not accounted for. Second, their data were analyzed using conventional Cox models, which are carried out under the assumption of noninformative censoring.^62^ However, in the presence of ≥1 competing risk,^63^ this assumption is violated, and can result in upward-biased effect estimates.^62, 64^ In this study, we accounted for a more robust set of confounders and used cause-specific Cox models to appropriately address competing risks, which may explain our comparatively lower adjusted HRs for both incident AFib and all-cause mortality.

Robust evidence suggests an association between history of HDP and risk of both CVD and mortality within the first decade following an affected pregnancy.^65, 66^ A recently published study^65^ reported a 1.2-fold increased risk of all-cause death in women <50 years of age with a history of any HDP compared to their unexposed counterparts. Despite this, only one published study^14^ reporting estimates of AFib risk in people exposed to HDP acknowledged the omission of mortality assessment as a study limitation.

In alignment with previous work,^65^ we found that a history of any HDP in a first obstetrical delivery was associated with a 30% increased rate of death without a prior AFib diagnosis after 9.5 years of follow-up. Importantly, the median age of subjects who died during the follow-up period was 29 years, with 67% of subjects being younger than 40 years of age when they died. Further, although both incident AFib and death were rare outcomes, death was slightly more common than AFib (2,951 vs. 2,483 respectively). These findings underscore the importance of considering mortality as a competing risk for non-fatal CVD outcomes in this relatively young population, as failure to do so could result in artificially inflated estimates of association.

We identified associations suggestive of dose-response relationships whereby both HDP severity and the presence of pre-pregnancy chronic hypertension were associated with higher cause-specific rates of both AFib and all-cause mortality. While the increasing magnitude of these point estimates suggests a dose-response pattern, cautious interpretation is needed due to overlapping 95% confidence intervals and non-significant estimates for specific diagnoses, likely due at least in part to insufficient power to examine specific associations among subtypes. Further, these associations did not directly align with our ‘hierarchical’ definition of HDP severity, which was assessed from the viewpoint of their acute clinical consequences for mother and fetus.^7^ Specifically, during exposure classification, we considered chronic hypertension in pregnancy to be the second most severe HDP diagnosis. However, it was associated with the highest csHazard of AFib and second highest csHazard of mortality without a prior AFib diagnosis [adjusted csHR (95% CI) 2.24 (1.55-3.23) and 2.04 (1.43-2.91), respectively]. Indeed, if future studies assessing CVD risk in people with a history of HDP continue to examine HDP as a composite exposure or adopt a narrow definition of HDP severity, critical dose-response associations may remain unidentified.^7^

Despite the obvious time-varying nature of HDP and CVD risk factors, most prior work assessing CVD risk in people with a history of HDP has employed a first pregnancy, time-fixed, approach to exposure ascertainment and data analysis.^67^ In this study, we also defined HDP exposure status according to subjects’ first delivery, and only adjusted for confounders that were assessed prior to cohort entry. Consequently, we were unable to account for changes in HDP exposure and confounder status over time and did not assess the cumulative effect of exposure to HDP in subsequent pregnancies on incident AFib. As such, our effect estimates may be subject to further residual confounding and/or other time-related bias.^68^ However, a few published studies, described as follows, can provide important insight about the robustness of this approach within our context.

One recent study^15^ reported that the adjusted hazard of arrhythmia for people with a history of preeclampsia in a first pregnancy was consistent with findings from a sensitivity analysis wherein preeclampsia was treated a time-dependent variable [adjusted HR (95% CI): 1.37 (1.23 to 1.54) vs. 1.29 (1.14 to 1.45), respectively]. Additional studies^68, 69^ have also found minimal differences between estimates of CVD risk generated using marginal structural (time-varying) Cox models compared to those generated using time-fixed analyses. As such, while the robustness of our findings should be assessed in models that account for the time-varying nature of HDP and confounders, prior research suggests that CVD risk may be primarily driven by HDP exposure in a first pregnancy and time-varying confounding may have a minimal impact on effect estimates.^68^

### Strengths and Limitations

Our large sample size and use of population-based data sources from the most populous ethnically and culturally diverse province in Canada enhances the generalizability of our findings. We followed established guidance on the reporting of studies carried out using routinely collected data,^26^ controlled for several important confounders, accounted for the competing risk of death, and our findings were robust to several sensitivity analyses.

Although we controlled for a robust set of confounders, we cannot rule out the possibility that residual confounding may have impacted our results. Pre-pregnancy BMI was unavailable for nearly 70% of subjects, thus, we adjusted for clinically documented obesity by identifying ICD-10 codes for this diagnosis in outpatient or inpatient records. However, individuals with codes for this condition likely only represent extreme cases, so control for this factor was likely incomplete, and may have resulted in spuriously inflated associations. Despite this, the results of our E-value sensitivity analysis provide some reassurance about the robustness of our findings to unmeasured confounding.

We considered incident chronic hypertension as an intermediate factor in the causal pathway between HDP and incident AFib and, thus, did not include it in multivariable models to prevent overadjustment bias.^70^ However, given the strong association between history of chronic hypertension in pregnancy and incident AFib identified in this study, the shared pathological features of chronic hypertension and AFib (e.g., left ventricular hypertrophy, diastolic dysfunction),^53^ and that incident chronic hypertension post-HDP was especially prevalent in subjects diagnosed with gestational hypertension (16%) and preeclampsia (14%), a formal causal mediation analysis^71, 72^ should be conducted to further elucidate the role that post-pregnancy chronic hypertension might play in this association.

## Conclusions

A history of gestational hypertension, preeclampsia, and chronic hypertension in pregnancy are each independently associated with a higher csHazard of incident AFib, with associations observed in relatively young women. Given its rapidly increasing global prevalence and significant morbidity and mortality burden, AFib has become one of the most substantial CVD challenges of the 21^st^ century– particularly for women. These findings underscore the critical importance of (a) incorporating information about history of HDP in risk calculation/stratification for both arrhythmic *and* non- arrhythmic forms of CVD, (b) improved population-based surveillance of traditional and female-specific CVD risk factors, and (c) the development of targeted prevention strategies designed to reduce the occurrence, and overall burden, of HDP.

## Acknowledgements

Parts of this material are based on data and/or information compiled and provided by CIHI, the Ontario Ministry of Health (MOH), and Immigration, Refugees and Citizenship Canada (IRCC) current to September 2020. The analyses, conclusions, opinions and statements expressed herein are solely those of the authors and do not reflect those of the funding or data sources; no endorsement is intended or should be inferred. This document used data adapted from the Statistics Canada Postal Code^OM^ Conversion File, which is based on data licensed from Canada Post Corporation, and/or data adapted from the Ontario Ministry of Health Postal Code Conversion File, which contains data copied under license from ©Canada Post Corporation and Statistics Canada. Adapted from Statistics Canada, this study used census 2001, 2006, and 2016. This does not constitute an endorsement by Statistics Canada of this product. We thank the Toronto Community Health Profiles Partnership for providing access to the Ontario Marginalization Index.

## Supplemental Material

Data Supplement S1-S8

Tables S1–S7

Figures S1-S5

References 1-22

